# Landscape of human gut antibiotic resistome and progression of diabetes

**DOI:** 10.1101/2021.06.24.21259507

**Authors:** Menglei Shuai, Guoqing Zhang, Fang-fang Zeng, Yuanqing Fu, Xinxiu Liang, Ling Yuan, Fengzhe Xu, Wanglong Gou, Zelei Miao, Zengliang Jiang, Jia-ting Wang, Lai-bao Zhuo, Yu-ming Chen, Feng Ju, Ju-Sheng Zheng

## Abstract

**Objective:** To investigate the association between human gut antibiotic resistome and the progression of type 2 diabetes (T2D) in a large cohort study.

**Design:** The present study included 1210 participants from the Guangzhou Nutrition and Health Study. We depicted the landscape of human gut antibiotic resistome with shotgun metagenomic data and examined its association with T2D and cardiometabolic traits. The co-occurrence network analysis was used to explore the associations between T2D-related ARGs and gut microbial species. We also examined the associations between gut antibiotic resistome features and fecal metabolome.

**Results:** There was a significant overall shift in gut antibiotic resistome structure among healthy, prediabetes and T2D groups (*p* = 0.004). We found that larger ARG diversity was associated with a higher risk of T2D (all *p* < 0.05). Based on the association found between resistomes and T2D, we developed diabetes ARG score and demonstrated its creative use as a new predictor for T2D progression manifested by the change of insulin resistance. Further network analysis showed the co-occurrence association between T2D-related ARGs and gut microbial species, which indicated the potential bacterial hosts of these ARG biomarkers. Resistome-metabolome co-analysis suggests a potential link of ARGs with fecal metabolites, which may reflect the host-microbial metabolic adaptation.

**Conclusion:** Our data depict the landscape of gut antibiotic resistome diversity and composition and uncover a close relationship between human gut antibiotic resistome and T2D progression.

Significance of this study

What is already known about this subject?

- Human gut is the key reservoir of microbiota harboring both commensal and pathogenic bacteria.
- Growing evidence has revealed that the gut microbiota composition was associated with type 2 diabetes (T2D).
- Prospective cohort studies showed that self-reported long-term use of antibiotics was associated with higher T2D risk.

What are the new findings?

- There was a significant overall shift in gut antibiotic resistome structure among healthy, prediabetes and T2D groups.
- We found for the first time that higher gut ARG diversity was associated with a higher risk of T2D.
- We constructed a diabetes ARG score (DAS) using the identified T2D related ARGs, it was positively associated with glycemic traits.
- Longitudinal validation analysis confirmed that the DAS was associated with T2D progression, characterized by the change of insulin resistance.

How might it impact on clinical practice in the foreseeable future?

- Our findings support a recommendation to avoid the abuse of antibiotics, and the identified antibiotic resistome markers could also be served as potential intervention target in future.

## Introduction

The global rise of antibiotic resistance jeopardizes the success and sustainability of modern medication to fight against deadly multi-drug resistant infections.^1^ Human gut is the key reservoir of microbiota harboring both commensal and pathogenic bacteria.^2^ It, however, also represents a hotspot for the frequent selection of diverse antibiotic resistance genes (ARGs) and their bacterial hosts due to the regular exposure of gut microbiota to foreign antibiotics directly from human medications and indirectly from food chain.^3,4^ While current cohort studies have largely focused on the microbiome diversity and its association with human disease and health, the research on gut antibiotic resistome, i.e., the collection of ARGs within microbiome, is rare in large human cohorts. The antibiotic exposure usually leads to long-term enrichment of ARGs in gut microorganisms,^5-7^ but its consequence and relation with human metabolic health were not clear.

We generated a hypothesis that gut antibiotic resistome may be associated with type 2 diabetes (T2D) progression, given that prior evidence suggested a close link between human gut microbiome and T2D,^8^ and that self-reported long-term use of antibiotics was associated with higher T2D risk and future cardiovascular diseases.^9,10^ Moreover, due to the genetic and lifestyle heterogeneities across different populations, we argued that compared with the prescriptions and questionnaires for inquiring antibiotic use, tracking the antibiotic resistome of gut microbiome would be more informative and help provide more mechanistic insight.

Therefore, the aim of the present study was to depict the landscape of gut antibiotic resistome in a large human cohort and explore its relationship with gut microbial metabolites and host metabolic health, which is a key step to understand the influence of the gut antibiotic resistome on human health.

## Methods

### Study design and participants

The current study was based on the Guangzhou Nutrition and Health Study (GNHS) which is an ongoing community-based cohort study in China. Between 2008-2013, a total of 4048 participants aged between 45 and 70 years, who lived in Guangzhou city for at least 5 years were enrolled in the GNHS. There were two waves for recruitment, 3169 were recruited between 2008 and 2010, and 879 were recruited between 2012 and 2013. All participants were followed up approximately every 3 years. Detailed information on the study design has been reported previously.^11^ The study was registered at clinicaltrials.gov (NCT03179657).

In the present study, participants were excluded according to the criteria: without metagenomics data (n = 2829), with missing data on main covariates including age, sex, BMI, smoking status, alcohol drinking, education, and income level (n = 2) or without Bristol stool scale information (n = 1). We also excluded participants who used antibiotics within two weeks before fecal samples collection (n = 6). Finally, 1210 participants were included in our analysis (Fig. S1). The study protocol was approved by the Ethics Committee of the School of Public Health at Sun Yat-sen University and Ethics Committee of Westlake University. All participants provided written informed consent.

### Metadata collection

#### Assessment of diabetes status and definitions

Diabetes status was defined according to the criteria of the American Diabetes Association and the World Health Organization: prediabetes was ascertained if a participant met one of the following criteria: (i) without a history of diabetes, (ii) glycated hemoglobin (HbA1c): 5.7%-6.4%, (iii) fasting blood glucose (FBG): 6.1-6.9 mmol/L; T2D cases were defined in adults as meeting one of the criteria: FBG ≥ 7.0 mmol/L, HbA1c ≥ 6.5% or self-reported medical treatment for diabetes.^12,13^

#### Covariates assessment

We collected the metadata on demographics, lifestyle, medical history and physical activity by questionnaires. Education attainment was categorized into primary (0-6 years), secondary (7-9 years) and higher education (≥ 10 years). Smoking status was categorized into current smoker and non-smoker. Alcohol drinking was classified as current drinker and non-drinker. Habitual dietary intakes at baseline were estimated from a validated food frequency questionnaire (FFQ), which recorded the frequencies of foods in the past 12 months. We then divided the food intakes into 16 food groups according to the Guidelines for Measuring Household and Individual Dietary Diversity.^14^

Physical activity was assessed as total metabolic equivalent for task (MET) hours per day based on a validated physical activity questionnaire, and it was classified into four groups according to quartiles. Body weight, height, waist circumference, hip circumference, neck circumference and blood pressure were measured by trained nurses on site. Fasting venous blood samples were collected at both baseline and follow-up visits. Glucose, total triglycerides, high-density lipoprotein (HDL) cholesterol, low-density lipoprotein (LDL) cholesterol, and total cholesterol in serum were measured on an automated analyzer (Roche cobas 8000 c702, Shanghai, China). Glycated hemoglobin (HbA1c) was measured with the Bole D-10 Hemoglobin A1c Program on a Bole D-10 Hemoglobin Testing System. Insulin was measured using the electrochemiluminescence immunoassay (ECLIA, Roche cobas 8000 e602) method. The homeostatic model assessment of insulin resistance (HOMA-IR) was calculated as fasting blood glucose (mmol/L) times fasting insulin (mIU/L) divided by 22.5.^15^

#### Fecal metagenomics profiling

Fecal samples of 1210 participants were collected during on-site study visits between 2015-2019. Before DNA extraction, the fecal samples were kept frozen at -80 °C. Fecal DNA extractions were carried out by a standardized CTAB procedure. DNA concentration was measured using Qubit dsDNA Assay Kit in Qubit 2.0 Fluorometer (Life Technologies, CA, USA). For DNA library preparation, a total amount of 1μg DNA per sample was used as input material. In addition, the NEBNext Ultra DNA Library Prep Kit (NEB, USA) was used following manufacturer’s recommendations and index codes were added to attribute sequences to each sample. The DNA samples were fragmented by sonication to a size of approximately 350 bp. Then, the DNA fragments were end-polished, A-tailed, and ligated with the full-length adaptor for Illumina sequencing with further PCR amplification. After that, PCR products were purified (AMPure XP system) and libraries were analyzed for size distribution by Agilent2100 Bioanalyzer and quantified using real-time PCR. The clustering of the index-coded samples was performed on a cBot Cluster Generation System according to the manufacturer’s instructions. After cluster generation, the library preparations were sequenced on an Illumina HiSeq platform and 150 bp paired-end reads were generated. Finally, we obtained on average 42.4 million paired-end raw reads for each sample (Table S3).

Next, raw sequencing reads were first quality-controlled with PRINSEQ (v0.20.4): 1) trim the reads by quality score from the 5′ end and 3′ end with a quality threshold of 20; 2) remove read pairs when either read was < 60 bp, contained “N” bases or quality score mean bellow 30; and 3) deduplicate the reads. Reads that could be aligned to the human genome (H. sapiens, UCSC hg19) were removed (aligned with Bowtie2 v2.2.5 using --reorder --no-contain --dovetail).^16^

Taxonomic profiling of the shotgun metagenomic data was performed using MetaPhlAn2 (v2.6.02), which uses a library of clade-specific markers to provide pan-microbial quantification at the species level.^17^ MetaPhlAn2 was run using default settings. Only species-level relative abundance data were considered in this study. Species were filtered out if their mean relative abundance and prevalence were <0.01% and <10% (Fig S2). Functional profiling was performed with HUMAnN2 v2.8.1, which maps sample reads against the sample-specific reference database to quantify gene presence and abundance in a species-stratified manner, with unmapped reads further used in a translated search against Uniref90 to include taxonomically unclassified but functionally distinct gene family abundances.^18^ Microbial gene richness (MGR) was computed by the number of genes present in each sample.

The ARG types and subtypes were annotated by ARGs-OAP v2.0 with default parameters.^19^ The expanded SARG database of ARGs-OAP v2.0 contains sequences not only from CARD and ARDB databases, but also carefully selected and curated sequences from the latest protein collection of the NCBI-NR database, to keep up to date with the increasing number of ARG deposited sequences. Each reference sequence is tagged with its functional gene annotation (ARG subtype) and membership within a class of antibiotics targeted by the gene (ARG type). For example, the prefix of *Tetracycline_TetQ* is ARG type and the suffix ‘*TetQ*’ is ARG subtype. The pipeline provides an algorithm for estimating cell number. The abundance of ARG was calculated and normalized by the cell number in our research, expressed as ‘copies of ARG per prokaryote’s cell’ using following equation:

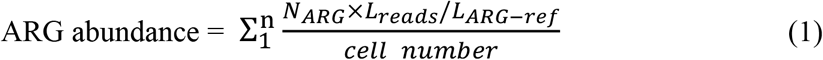

Where n is the number of ARG reference sequences belonging to an ARG type or subtype; *N*_*ARG*_ is the number of the ARG-like sequences annotated to one specific ARG reference sequence in the metagenome; *L*_*reads*_ is the sequencing length of metagenomic reads; *L*_*ARG-ref*_ is the average length of the correspondingly specific ARG reference sequences. The cell number was computed based on the following equation^20^:

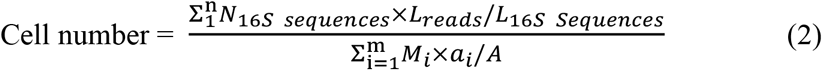

Where *m* is the total taxa detected from the metagenomics dataset based on the extracted hyper variable region information; *M*_*i*_ represents the number of copies of taxon *i* from the CopyRighter database; *a*_*i*_ is the number of aligned hypervariable sequences of taxon *i* in the metagenomics datas set; *A* is the total number of aligned hypervariable sequences in all *m* taxa.

In addition, we excluded the ARG types or subtypes that were present in less than 10% of the samples (Fig S2). Unclassified ARG types or subtypes were also excluded in current study. The α-diversity of gut antibiotic resistome was represented by three diversity indices: Shannon, Richness (Observed unique ARGs) and Evenness (Pielou’s index), which was estimated by the *vegan* R package.^21^ Details of genotyping data processing, targeted fecal metabolome profiling and statistical analyses were described in online supplementary materials.

## Results

### Composition and variation of the gut antibiotic resistome

We included 1210 participants with a mean age of 64.9 years (SD: 5.5) and with fecal metagenomics data from the GNHS into our present analysis (Fig. 1).^22^ We assessed the diabetes status of the participants, of whom 531 were healthy, 495 were prediabetes and 184 were T2D (table 1). Using paired-end shotgun metagenomic sequencing, we obtained 42.4 million paired-end reads on average (Table S2).

**Fig. 1.**
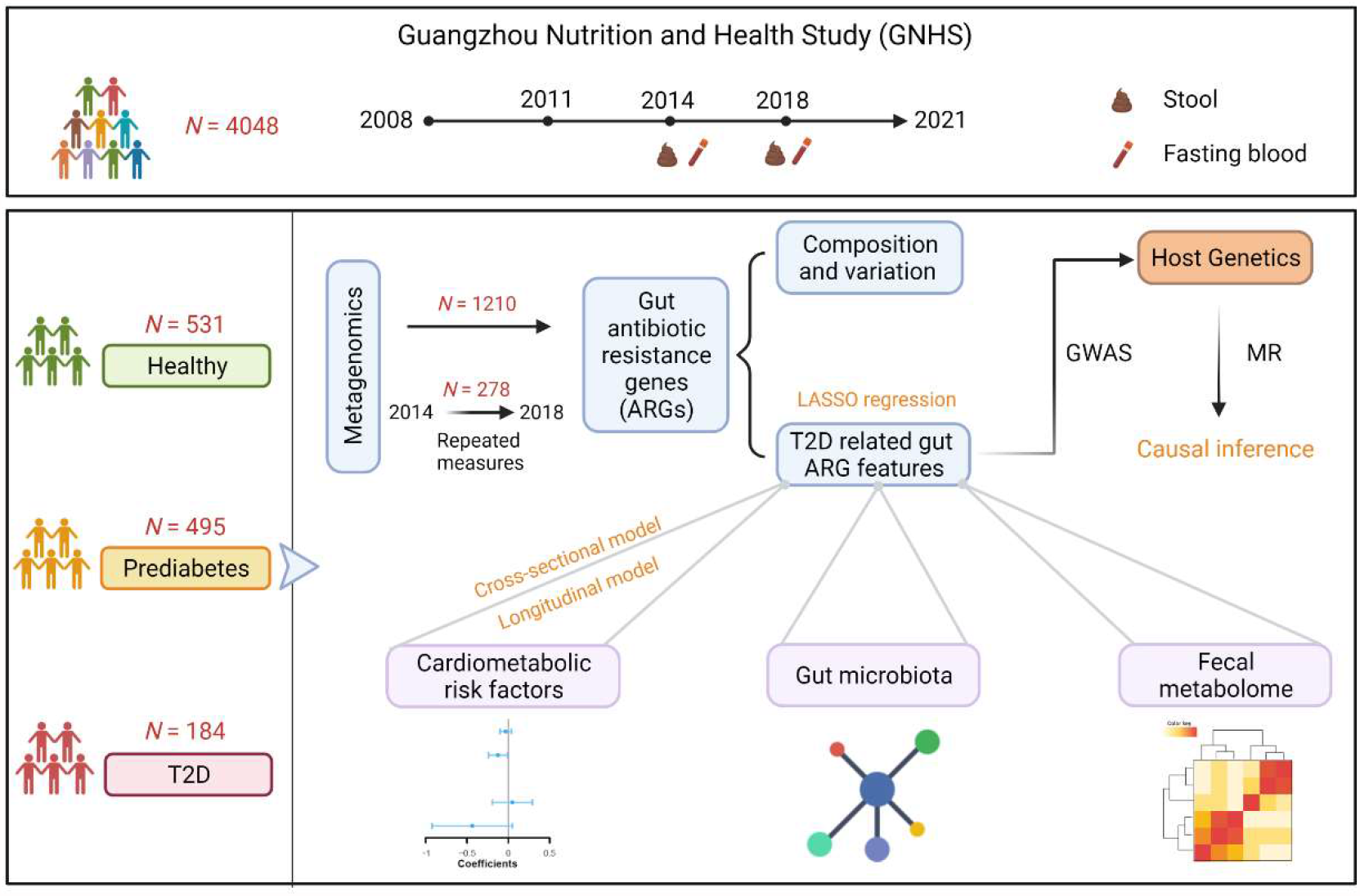
Study strategy for linking gut antibiotic resistome and progression of T2D. Our study was based on the Guangzhou Nutrition and Health Study (GNHS) which is an ongoing community-based cohort study in China. We included 1210 participants in the current study, of whom 531 were healthy, 495 were prediabetes and 184 were T2D. To associate the gut antibiotic resistome and T2D, we profiled the stool metagenome. 278 individuals have collected stool sample twice at different time points. We applied ARG-OAP2 and MetaPhlAn2 to perform the profiling of the gut antibiotic resistome and gut microbiota, respectively. To explore the potential causal association of the identified ARG features with T2D, we performed genome-wide association analyses (GWAS) and one-sample Mendelian randomization (MR) analyses. In addition, we examined the associations between gut ARG features and cardiometabolic traits using the cross-sectional model, which was validated by the longitudinal model. We also constructed the co-occurrence network between gut ARGs and microbial species. Finally, Spearman correlation analysis was used to investigate the associations between T2D related ARG features and fecal metabolome.

We detected a total of 19 ARG types and 805 ARG subtypes (Fig. S2). The most abundant ARG types in this study included *Multidrug, Tetracycline* and *Macrolide-Lincosamide-Streptogramin (MLS)*, followed by *Beta-lactam, Aminoglycoside* and *Bacitracin* (Fig. 2A). After 10% prevalence filtering, 17 ARG types and 233 subtypes remained for further analysis. We observed that *Tetracycline*_*TetQ* was the most abundant ARG subtype in our populations (Fig. 2B). The prevalence of *Quinolone, Quinolone_norB* and subtypes of *Beta-lactam* increased along with the diabetes progression (Fig. S3A-C). Overall, 17 core ARG subtypes were shared by all of the participants among healthy, prediabetes and T2D groups (Fig. S3D).

**Fig. 2.**
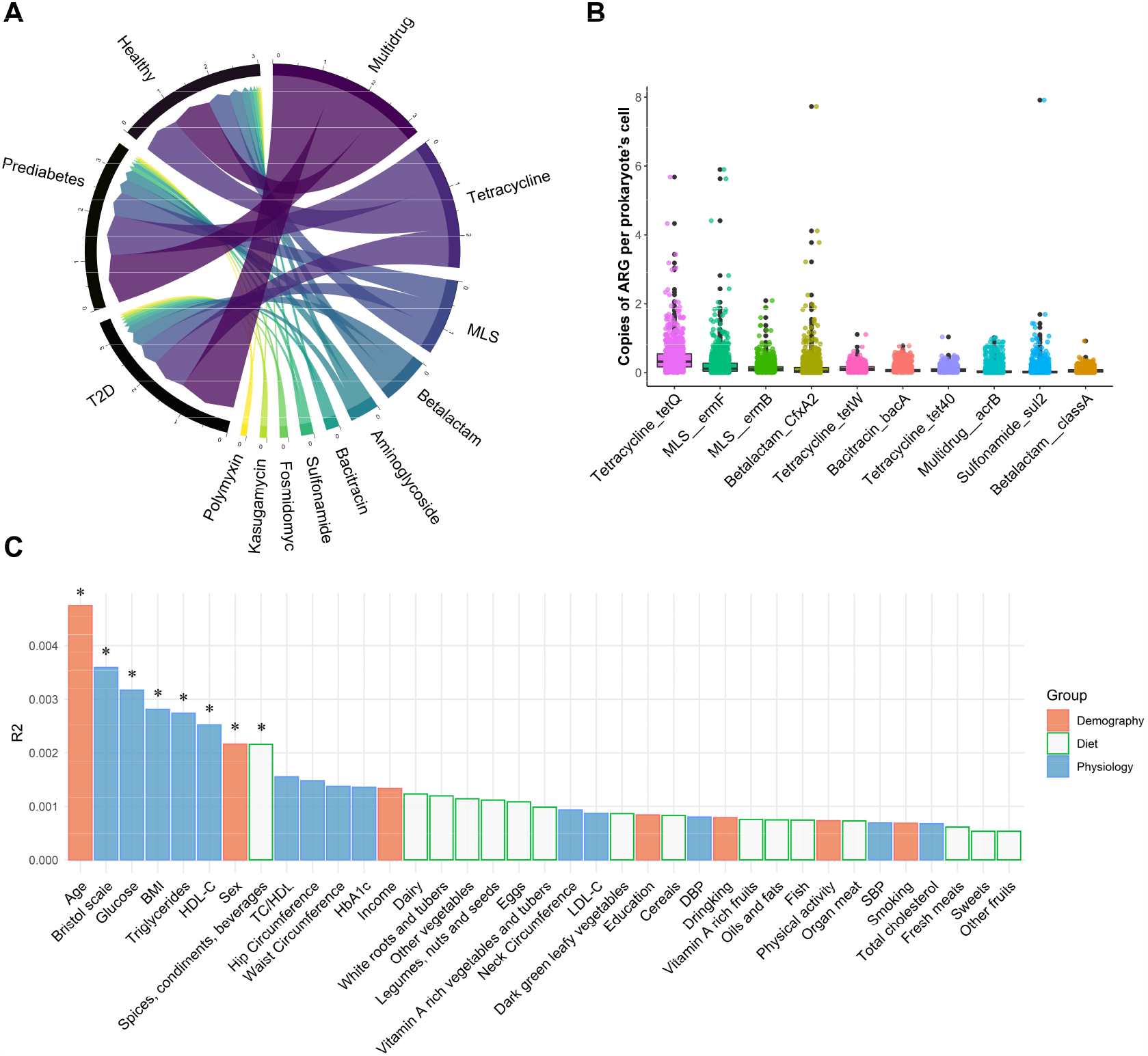
Compositional variation in human gut antibiotic resistome. **A**, Circus plot showing the mean relative abundance of top 10 antibiotic resistance genes (ARGs) types among Healthy (n = 531), Prediabetes (n = 495) and Type 2 Diabetes (T2D, n = 184) groups. **B**, Box plot showing the abundance of top 10 ARGs subtypes (n = 1210). **C**, The effect sizes of host factors on human gut antibiotic resistome were calculated by PERMANOVA (Adonis, permutations = 999) (n = 947). MLS, Macrolide-Lincosamide-Streptogramin.

We correlated 7 demographic factors, 16 dietary intakes and 14 physiological factors to the inter-individual variation in the gut antibiotic resistome. Age had the largest explanatory power on the gut antibiotic resistome compositional difference (Bray-Curtis distance, PERMANOVA, *p* = 0.002), which may be due to the accumulation of ARGs during the lifespan and become more diversified with age.^23^ A total of 8 factors were found to be significantly associated with the overall resistome variation (Fig. 2C and Table S3). In addition, we repeated our analysis among healthy, prediabetes and T2D groups, respectively. Our data showed different patterns for the contribution of those factors, which indicates the impact of diabetes status on the relationship between gut antibiotic resistome and those factors (Fig. S4).

### Gut antibiotic resistome composition are associated with type 2 diabetes

In total, 639 bacterial species were identified in the 1210 samples. To focus on more representative species, 156 were kept for downstream analysis after filtering by relative abundance (mean> 0.01%) and prevalence (>10%) (Fig. S2 and Table S11). Pearson’s correlation analyses indicated a positive association between α-diversity of the gut antibiotic resistome and microbial gene richness (MGR) which is an α-diversity indicator of the gut microbiota (*r* = 0.27-0.29, *p* < 2.2×10^−16^) (Fig. S5A-B). Similarly, Procrustes analysis demonstrated a strong cooperativity of the gut antibiotic resistome and gut microbiota profiles (Fig. S5C). Notably, our data showed that MGR was inversely associated with prevalent T2D after multivariable adjustment (Odds Ratio (OR) = 0.67, *p* < 0.001) (Fig. 3C). Therefore, we further performed a multivariable logistic regression analysis to investigate the association between α-diversity of the gut antibiotic resistome and T2D, adjusted for MGR and other potential confounders. We found that larger Shannon index (OR = 1.20, 95% CI 1.01-1.41, *p* = 0.034) and observed richness (OR = 1.20, 95% CI 1.02-1.42, *p* = 0.029) of ARGs were associated with a higher risk of T2D (Fig. 3C). Moreover, there were significant shifts for the gut antibiotic resistome composition across the healthy, prediabetes and T2D individuals (Bray-Curtis distance, PERMANOVA, *p* = 0.004) (Fig. 3A, B and Fig. S6). Interestingly, we observed a significant difference in the composition of gut ARG subtypes (*p* = 0.029) between healthy and prediabetes participants, but no significant change in the gut microbiota composition between the two groups (*p* = 0.304) (Table S4).

**Fig 3.**
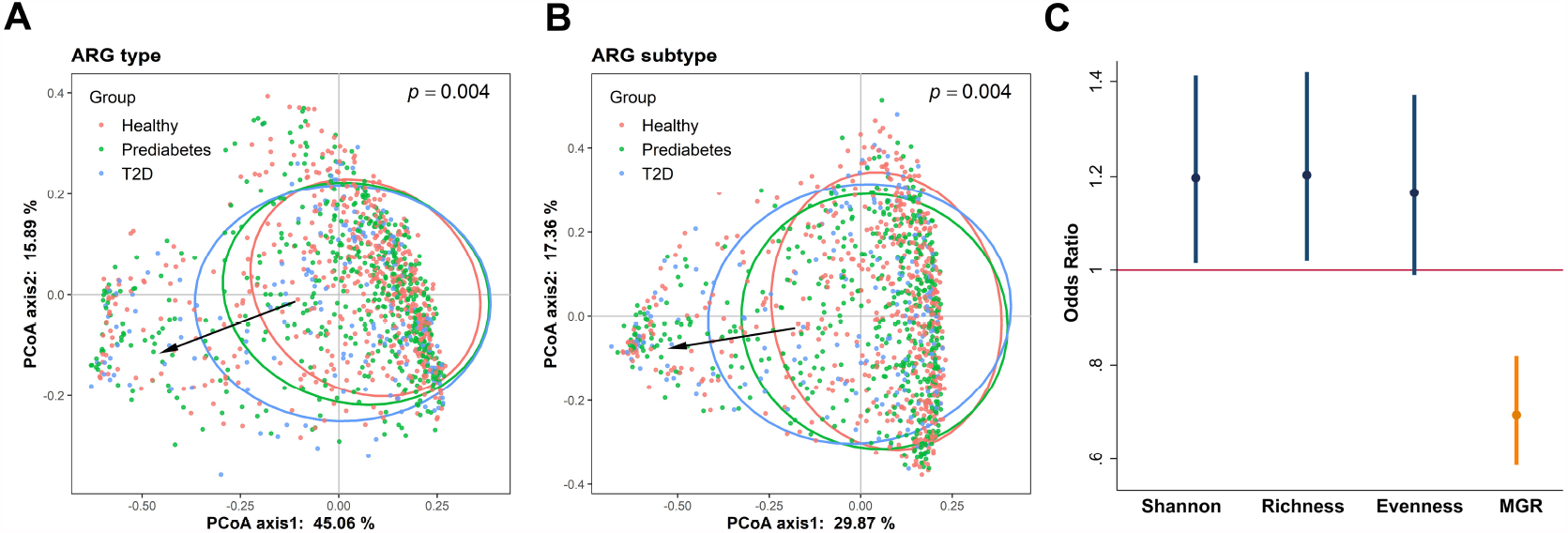
Gut antibiotic resistance genes are associated with type 2 diabetes. **A** & **B**, PCoA plots showing the compositional differences of gut antibiotic resistome among different diabetes groups. *p* was calculated by PERMANOVA (Adonis, permutations = 999) based on Bray-Curtis distance. **C**, Forest plot showing the odds ratio of T2D for each metagenomics diversity index (in SD unit). Logistic regression models were adjusted for age, sex, BMI, smoking status, drinking status, education attainment, household income level, physical activity, Bristol stool scale, and microbial gene richness (MGR). The error bars represent confidence intervals. * *p* < 0.05.

### Shifts in gut antibiotic resistome and microbial species with diabetes status

We then identified 25 ARGs and 27 microbial species related to T2D, based on the least absolute shrinkage and selection operator (LASSO) regression model (Fig. 4A and Table S5, S6).^24^ Considering the progression effects of T2D, we combined the selected features from three binary dependent variable models: non-T2D (healthy and prediabetes)/ T2D, healthy/T2D, and prediabetes/T2D (Fig. S7, S8). Among those markers, we observed a different overall pattern shift in the abundance for gut ARGs and microbial species from healthy, prediabetes to T2D. Specifically, the changes of abundance in 25 ARGs were divided into three distinct clusters while 27 microbial species were divided into two clusters (Table S7). Among the ARGs, *Vancomycin_vanX, Multidrug_emrE, MLS_ermX* and *Quinolone_norB* were positively associated with T2D risk (OR = 1.15-1.18, *p* < 0.05) (Fig. 4A). We then used a one-sample Mendelian randomization (MR) analyses to explore the potential causal association of the above identified ARG features with T2D. To enable the MR study, we performed genome-wide association analyses for the selected ARG features and constructed genetic instruments for these features (Table S8). We found that genetically predicted higher abundances of *Multidrug_emrE* and *MLS_ermX* were associated with higher T2D risk (OR = 1.10, 95% CI 1.04-1.16, *p* = 0.004; OR = 1.10, 95% CI 1.03-1.17, *p* = 0.01; Fig. 4B). We also observed that genetically predicted higher levels of gut ARG richness were associated with higher T2D risk (OR = 1.26, 95% CI 1.00-1.50, *p* = 0.07, Fig. 4B).

**Fig. 4.**
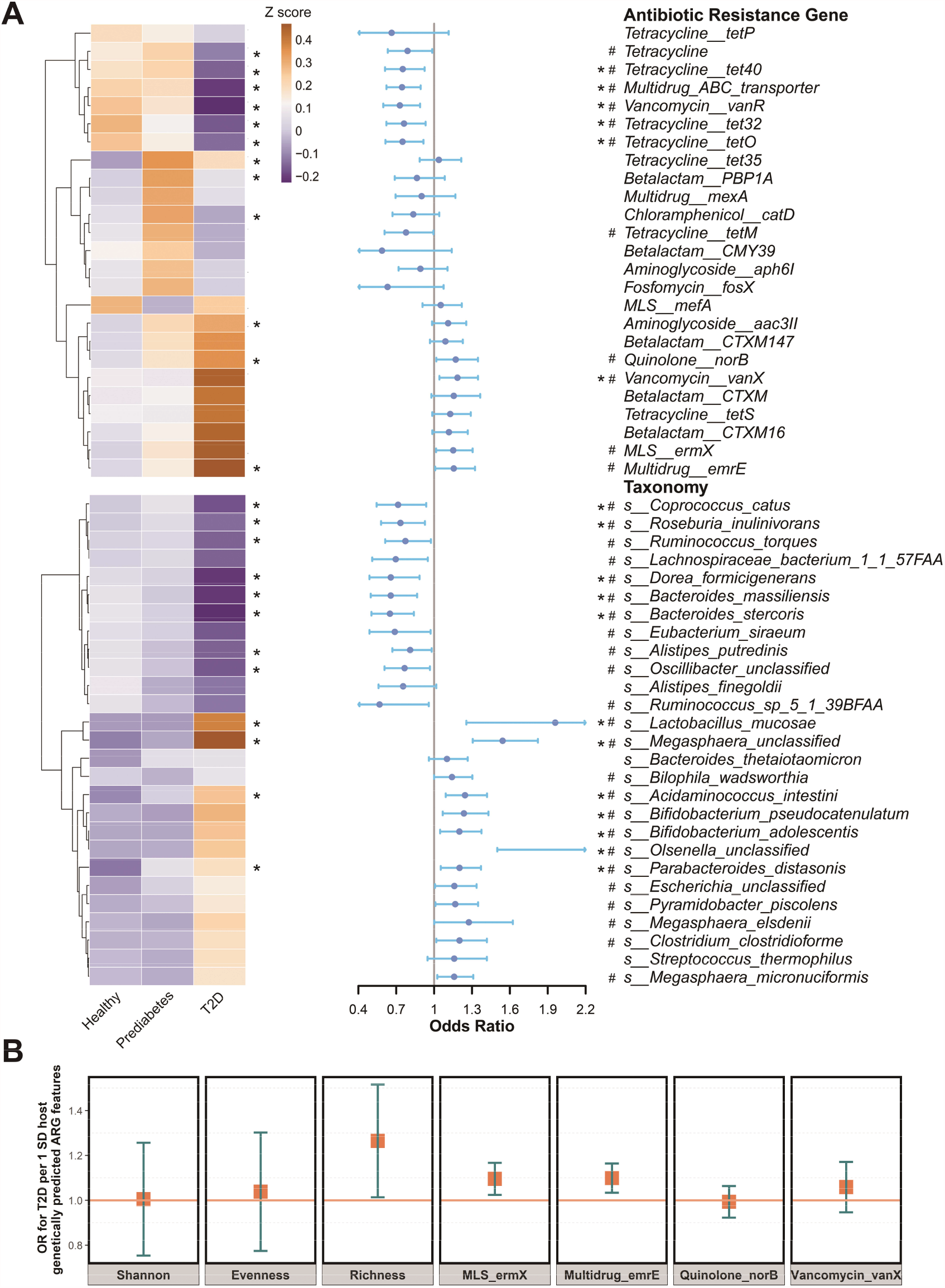
Shifts in gut antibiotic resistome and microbial species with diabetes status. **A**, Left heatmap shows the mean relative abundance (scaled by z score normalization) of the 25 identified T2D-related ARGs and 27 identified T2D-related microbial species from LASSO model. The abundance differences of these markers in healthy, prediabetes and T2D groups were examined by Kruskal–Wallis test. Right forest plot shows the odds ratio of T2D for each marker. Logistic regression models were performed, adjusted for age, sex, BMI, smoking status, drinking status, education attainment, income level and physical activity. The error bars represent confidence intervals. **B**, Mendelian randomization analysis (n = 947). Forest plot shows the odds ratios (95% confidence intervals) of T2D (outcome) per standard deviation increase in the host genetically predicted levels of the above identified ARG features (exposure). *p* values were corrected by Benjamini-Hochberg method. MLS, Macrolide-Lincosamide-Streptogramin. *FDR-corrected *p* < 0.05. #raw *p* < 0.05.

### Gut antibiotic resistome features are associated with cardiometabolic risk factors

Based on the collection of T2D-related gut ARGs, we constructed a novel Diabetes-ARG Score (DAS) which was positively associated with T2D (*p* = 7.8×10^−6^), to represent the T2D-related gut antibiotic resistome. We found that the DAS was significantly associated with glycemic traits, such as fasting blood glucose (FBG), glycated hemoglobin (HbA1c), insulin, and homeostatic model assessment of insulin resistance (HOMA-IR). For instance, the multivariable linear regression model indicated that a higher DAS was associated with a higher level of FBG and HOMA-IR (all *p* < 3.2×10^−4^) (Fig. 5A). Furthermore, we used a linear mixed model to investigate the longitudinal associations between DAS and glycemic traits after excluding the baseline T2D cases. Our analyses further validated the positive association of DAS with insulin and HOMA-IR (both *p* < 5×10^−3^), suggesting that baseline gut antibiotic resistome was prospectively associated with T2D progression, characterized by the alteration of insulin resistance (Fig. 5B).

**Fig. 5.**
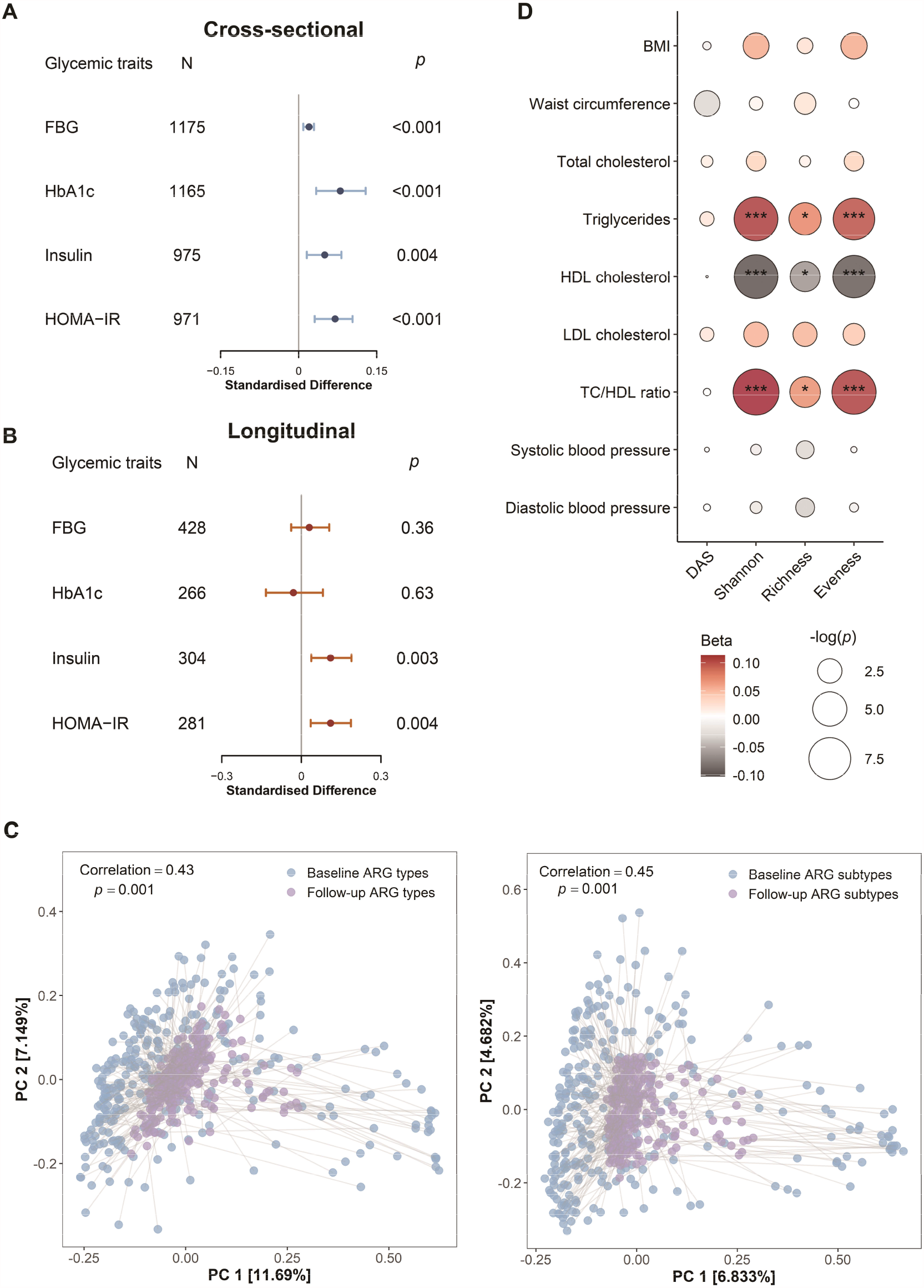
Gut antibiotic resistome features are associated with glycemic and cardiometabolic traits. **A**, Forest plot showing the cross-sectional associations between DAS and glycemic traits. Linear regression models were performed, adjusted for age, sex, BMI, smoking status, drinking status, education attainment, income level, physical activity and Diabetes-Microbiota Score (DMS). Standardized difference is the difference (in SD unit of the glycemic traits) per 1 SD change of DAS. The error bars represent confidence intervals. **B**, Forest plot showing the longitudinal associations between the baseline DAS and glycemic traits (repeated measured at baseline and a follow-up visit). Linear mixed models were performed, adjusted for potential covariates the same as above linear model. Standardized difference is the difference (in SD unit of the glycemic traits) per 1 SD change of DAS. The error bars represent confidence intervals. **C**, Procrustes analysis of gut ARGs at the baseline versus gut ARGs at a follow-up visit (n = 278). The median follow-up time was 3.2 years. Baseline and follow-up visit ARGs are shown as blue and purple dots, respectively. Baseline and follow-up visit ARGs from the same individual are connected by grey lines. **D**, Associations between gut antibiotic resistome features and cardiometabolic risk factors. Linear regression models were used, adjusted for potential covariates the same as above linear model. Total number of participants in each analysis was 1175 for HDL cholesterol, LDL cholesterol, total cholesterol and TC/HDL ratio, 1176 for triglycerides, 1210 for diastolic blood pressure, systolic blood pressure and BMI, and 1203 for waist circumference. DAS, Diabetes-ARG Score, FBG, insulin, HOMA-IR were log-transformed. FBG, fasting blood glucose, HbA1c, glycated hemoglobin; HOMA-IR, homeostatic model assessment of insulin resistance; BMI, body mass index; HDL, high-density lipoprotein; LDL, low-density lipoprotein. HbA1c, glycated hemoglobin. Triglycerides and TC/HDL ratio were log-transformed. \**p* < 0.05.

The Procrustes analysis in 278 participants of the cohort shows that the gut antibiotic resistome had a significant consistency between the two time points (baseline and a follow-up visit) (Fig. 5C). Moreover, we also observed associations between gut antibiotic resistome features and other cardiometabolic traits. The α-diversity indices were positively associated with triglycerides and total cholesterol/ high-density lipoprotein (HDL) cholesterol ratio, while inversely associated with HDL cholesterol (Fig. 5D and Table S10).

### Co-occurrence patterns among T2D-related ARGs and microbial species

The positive correlations between the T2D-related ARGs and microbial species suggest that the inverse correlations between ARGs and T2D may be caused by the depletion of beneficial bacteria. For instance, *Multidrug_ABC_transporter, Vancomycin_vanR* and *Tetracycline*_*tet32* were positively associated with *Roseburia inulinivorans* which is a producer of butyric acid (Spearman’s *rho* = 0.34, 0.25 and 0.3, FDR-corrected *p* < 1.9×10^−17^) (Fig. 6A and Table S12). A prior study showed that *R. inulinivorans* tended to be depleted in the gut microbiome of individuals with T2D, which was in line with our study (Fig. 4A).^8^ Moreover, co-occurrence network depicted the positive associations (Spearman r ≥ 0.3 and FDR-corrected *p* < 0.05) between the T2D-related ARGs and all of the bacteria species, which could be an effective way to track their potential hosts and co-occurring bacteria of human gut or environmental ARGs.^25-27^Specifically, both our data and the literature confirmed that *Escherichia* (s575 and s578) carries the *Multidrug_emrE* (Fig. 6B and Table S13-16).^28^ Here, T2D was first found to be positively associated with *Multidrug_emrE* (OR = 1.16, 95% CI 1.01-1.32, *p* = 0.037). Our data also showed that the number of edges of the ARG-Species association networks had a gradual increment from heathy (87) to prediabetes (100) and T2D (128), suggesting that gut ARGs might be observed in more bacteria species with the disease progression (Table S17). The above results together revealed a close relationship between the gut antibiotic resistome and T2D progression.

**Fig. 6.**
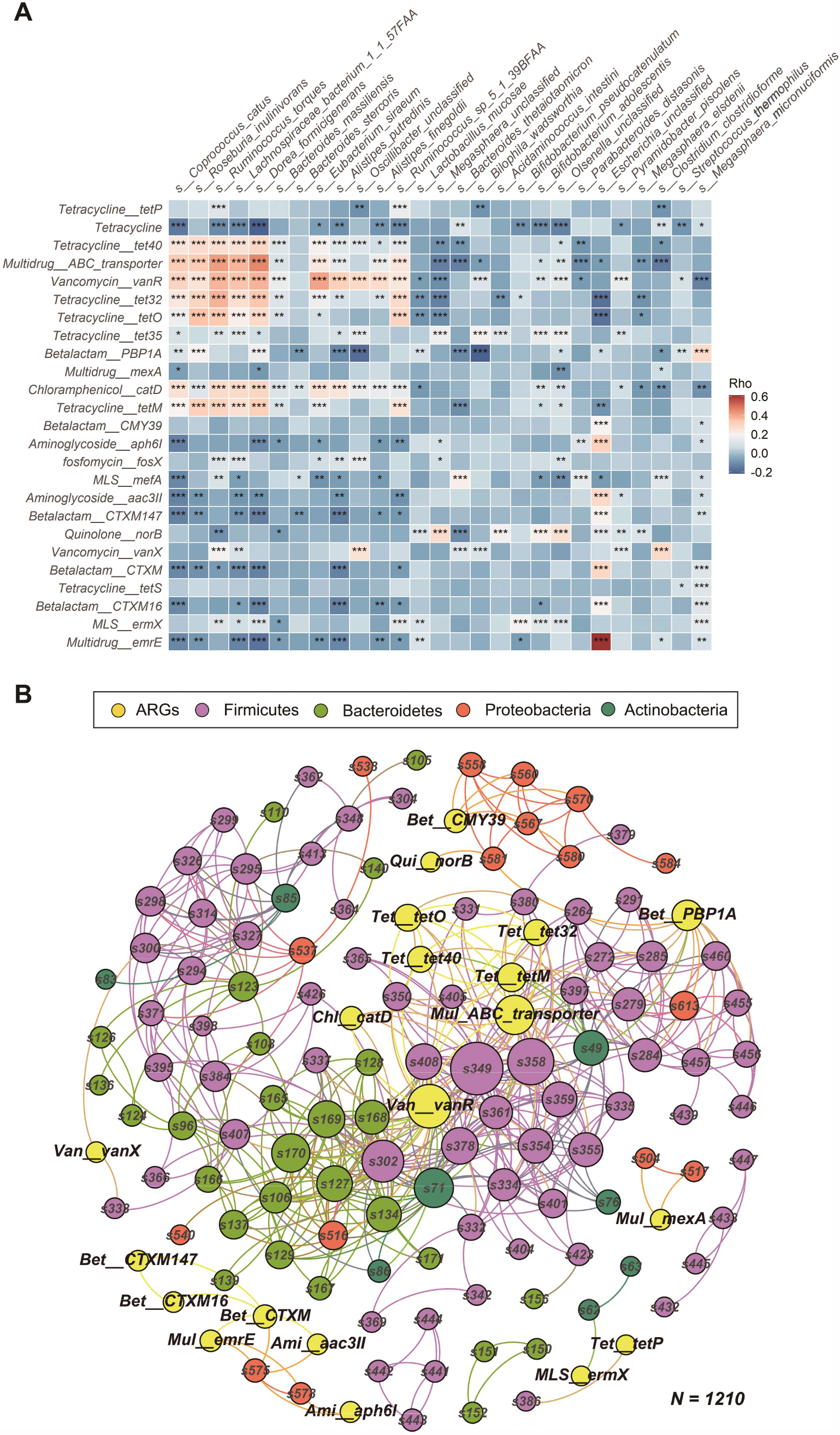
Co-occurrence patterns among T2D-related ARGs and microbial species. **A**, The heatmap shows the Spearman correlation coefficients between T2D-related gut antibiotic resistance genes (ARGs) and T2D-related microbial species. **B**, Correlation-based networks of co-occurring T2D-related ARGs and all microbial species colored by node affiliation. A node stands for an ARG type/subtype or a species and a connection (i.e. edge) stands for a significant (FDR-corrected *p* < 0.05, Spearman’s rho ≥ 0.3, n =1210) pairwise correlation. Node size is proportional to the number of connections (i.e., degree). Network was colored by ARGs and phylums. *Ami, Aminoglycoside*; Bet, *Betalactam*; *Chl, Chloramphenicol*; *MLS, Macrolide-Lincosamide-Streptogramin*; *Mul, Multidrug*; *Qui, Quinolone*; *Tet, Tetracycline*; *Van, Vancomycin*; *FDR-corrected *p* < 0.05, ** FDR-corrected *p* < 0.01, ***FDR-corrected *p* < 0.001.

Plasmid-mediated quinolone resistance in *Klebsiella pneumonia* was discovered in 1998, and it could transfer low-level quinolone resistance to other bacteria.^29^ Here, the *Quinolone_norB* was only significantly co-occurred with *Klebsiella pneumonia* (s581) in healthy and prediabetes populations (Fig. S9). However, in the T2D group this gene was also found to newly co-occur with an unclassified *Olsenella* species (s89) which had a positive association with T2D (Fig. 4A). In the global co-occurrence networks that contain all the ARGs and microbial species, we found that ARGs tended to co-occur more likely (O/R ratio = 1.67) with T2D-positively related microbial species (T2D-pos-Spe) whereas less likely (O/R ratio = 0.21) with T2D unrelated species (ARG-Non-T2D-Spe) than expected under random association (Table S18). In addition, the co-occurrence pattern between ARGs and T2D-pos-Spe tended to be randomized in prediabetes populations (as suggested by O/R ratio = 0.89), implying the potential for the spread of ARGs during the period of prediabetes, an early developmental stage of T2D.

### Gut antibiotic resistome features are associated with fecal metabolome

Antibiotic resistance in bacteria is often associated with a metabolic burden, while microbial metabolic adaptations accompanying the development of antibiotic resistome in T2D patients are unclear.^30,31^ We therefore examined the associations between gut antibiotic resistome features and 117 targeted fecal metabolites. As expected, the gut antibiotic resistome was widely associated with the fecal metabolites. Specifically, we found that DAS and *Vancomycin_vanX* were positively associated with L-isoleucine and L-leucine (all FDR-corrected *p* < 0.01) (Fig. S10). In addition, we observed that DAS and *Multidrug_emrE* were positively associated with 10-trans-heptadecenoic acid and 8,11,14-eicosatrienoic acid, while negatively associated with butyric acid (Fig. S10).

## Discussion

Here, we not only demonstrate the previously unperceived association between ARGs and microbiota, but also discover a shift and spread in gut antibiotic resistome with the progression of T2D, which may accompany with widespread host-microbial metabolic adaptations. Our study unravels a novel link between the gut microbiome and progression of T2D via antibiotic resistome.

A previous small-scale study showed that *TetQ* and *ErmB* were the top two abundant ARGs in the Chinese.^32^ Similarly, we observed that *Tetracycline_TetQ* was also the most abundant ARG subtype in our cohort participants, and *MLS_ermB* was the third abundant ARG subtype. Moreover, six of the top ten abundant ARG subtypes were consistent with the study aforementioned.

Notably, we found for the first time, to the best of our knowledge, that ARG diversity was positively associated with T2D. In contrast, previous studies showed that the diversity of gut microbiota is inversely associated with T2D.^33,34^ To avoid potential confounding effect of the gut microbiota, our model was adjusted for the total microbial gene richness. These results suggest that the diversity of the gut antibiotic resistome is different from that of the gut microbiota, in terms of their relationships with metabolic disease. In addition, we observed a significant difference in the composition of gut ARG subtypes between healthy and prediabetes participants, but no significant change in the gut microbiota composition between the two groups. The latter is in line with a previous study which reported no significant microbiota change between the impaired fasting glucose tolerance participants and the low-risk normal glucose tolerance participants^35^. These results suggest that the gut antibiotic resistome may change earlier than the gut microbiota during the progression of T2D, and/or that changes in the gut antibiotic resistome are more sensitive to the development of T2D.

Although it is hard to determine whether the gut ARG features causally increase the T2D risk, our data indirectly permit fairly speculation. For example, the results of MR analyses in our study reveal a potential causal links between gut ARG richness, *Multidrug_emrE* and T2D. *Multidrug_emrE* is a small-drug efflux pump, which confers resistance to a wide variety of antimicrobial agents including cationic disinfects (e.g., quaternary ammonium compounds used in the hospitals and food industry)^36^ and antibiotics (e.g., ampicillin, erythromycin, and tetracycline).^37^ Nevertheless, the current MR analyses were preliminary and exploratory, given the limited availability of genetic instrument for analyzing ARG features. More studies are required to further elucidate the causal links between gut ARGs and T2D.

Previous evidence showed that plasma L-isoleucine and L-leucine had highly significant associations with future diabetes risk^38^. In our study, both of them in fecal samples were positively associated with DAS and *Vancomycin_vanX*. Moreover, we observed that DAS and *Multidrug_emrE* were positively associated with 8,11,14-eicosatrienoic acid, which is also known as dihomo-gamma-linolenic acid (DGLA). A cross-sectional study revealed that the DGLA level was an independent determinant for HOMA-IR in 225 Japanese patients with T2D.^39^ Furthermore, our study found that the gut antibiotic resistome was widely associated with the fecal metabolites, which may reflect the host-microbial metabolic adaptation. Bacteria could develop resistance to many classes of antibiotics vertically, by making mutations in central housekeeping genes, which correspondingly affected metabolism.^40^ One study on *E. coli* demonstrated that the acquisition of antibiotic resistance is accompanied by specifically reorganized metabolic networks in order to circumvent metabolic costs.^41^ Taken together, these results provide a potential interpretation for the mechanism behind the observed association in the present study.

The present study has several strengths. First, we profile the gut antibiotic resistome configuration in a large population with different diabetes status, which has not been investigated before. Second, previous studies mainly focus on the relationship between gut microbiota and T2D, while we also examine the association between gut antibiotic resistome and T2D progression, and the results were validated by the longitudinal analyses. Finally, we modelled, for the first time, the association between the antibiotic resistance and T2D and constructed a novel diabetes-ARG score, which may help inform a new mechanism discovery for the pathophysiology of T2D. Nonetheless, there are several limitations. First, our study is based on the Chinese population and may not be generalizable to other ethnicities. Therefore, the generalization of our findings would require validation in other countries or ethnicities. Second, our results are from an observational study, which is subject to the influence of residual confounders. Third, detailed mechanism behind our observed association is not clear, more mechanistic investigation in future is needed to provide causality.

In conclusion, our study depicts a comprehensive landscape of the gut antibiotic resistome in a large human cohort and provides a novel insight about the relationship between antibiotic resistance and T2D progression. These results also suggest that the gut antibiotic resistome is closely connected with fecal metabolites and host metabolic health. These novel ARG features may potentially serve as intervention target of T2D in future studies.

## Data Availability

The raw data of metagenomic sequencing is available in the CNSA (https://db.cngb.org/cnsa/) of CNGBdb at accession number CNP0001510. Other data sets generated during and/or analyzed during the current study are available from the corresponding author upon reasonable request.

## Acknowledgements

We thank all study participants of the Guangzhou Nutrition and Health Study. We thank Westlake University Supercomputer Center for assistance in data storage and computation. We thank the support from Westlake Education Foundation.

## Author contributions

JSZ, FJ, YMC: designed research; CWL, FX: collected the data; MLS, GQZ, FJ and LY: performed the data analysis; MLS, FJ, JSZ: wrote the manuscript (MLS drafted the initial manuscript; JSZ, FJ, GQZ and MLS finalized the manuscript); JSZ, YMC, FJ, YQF, GQZ, XXL, ZM, ZJ: revised the manuscript; JSZ and FJ: had primary responsibility for the final content; All authors read, revised and approved the final manuscript.

## Code availability

The scripts and instructions used for metagenomics processing pipeline are available from https://github.com/emblab-westlake/rMAP. For ARGs annotation, we used the open-source ARG-OAP2 pipeline, which is available at https://github.com/biofuture/Ublastx_stageone.

## Patient and public involvement

Patients and/or the public were not involved in the design, or conduct, or reporting, or dissemination plans of this research.

## Abbreviations

ARG: antibiotic resistance genes;
T2D: type 2 diabetes;
rRNA: ribosomal RNA;
GNHS: Guangzhou Nutrition and Health Study;
FFQ: food-frequency questionnaire;
PCoA: principal coordinate analysis;
PERMANOVA: permutational multivariate analysis of variance;
TC: total cholesterol;
HDL: high-density lipoprotein;
LDL: low-density lipoprotein;
HbA1c: glycated haemoglobin;
DAS: Diabetes ARG score;
*MLS*: *Macrolide-Lincosamide-Streptogramin*;
HOMA-IR: homeostatic model assessment of insulin resistance

