## Supplementary Material for "Landscape of human gut antibiotic resistome and progression of diabetes"

**This word file includes:**

Materials and Methods

**Study design and participants**

**Metadata collection**

**Fecal metagenomics profiling**

**Genotyping data**

**Targeted fecal metabolome profiling**

**Statistical analysis**

Figs. S1 to S10

**Fig. S1. Flow diagram of participants' selection for the analyses of present study.**

**Fig. S2. Workflow for analysis of metagenomics sequencing data.**

**Fig. S3. Abundance and prevalence of gut antibiotic resistome in GNHS.**

**Fig. S4. The effect sizes of host factors in gut antibiotic resistome grouped by diabetes status.**

**Fig. S5. Correlations between gut antibiotic resistome diversity and microbiota.**

**Fig. S6. The comparison of inter-individual Bray-Curtis distance of gut antibiotic resistome.**

**Fig. S7. The performance of models based on LASSO feature selection.**

**Fig. S8. Veen plot of biomarkers identified by LASSO models.**

**Fig. S9. Networks of co-occurring T2D-related ARGs and gut microbiota, grouped by diabetes status.**

**Fig. S10. Associations between gut antibiotic resistome features and fecal metabolites (n = 1012).**

### Materials and Methods

#### Genotyping data

Host DNA was extracted from leukocytes using the TIANamp® Blood DNA Kit (DP348, TianGen Biotech Co, Ltd., China) according to the manufacturer's instructions. DNA concentrations were determined using the Qubit quantification system (Thermo Scientific, Wilmington, DE, USA). Extracted DNA was stored at −80 °C. Genotyping was carried out with Illumina ASA-750K arrays. Quality control and relatedness filters were performed by PLINK1.9 [1]. Individuals with a high or low proportion of heterozygous genotypes (outliers defined as 3 standard deviations) were excluded [2]. Individuals who had different ancestries (the first two principal components ± 5 standard deviations from the mean) or related individuals (IBD > 0.185) were excluded [2]. Variants were mapped to the 1000 Genomes Phase 3 v5 by SHAP EIT [3,4], and then we conducted genome-wide genotype imputation with the 1000 Genomes Phase 3 v5 reference panel by Minimac3 [5,6]. Genetic variants with imputation accuracy RSQR > 0.3 and MAF > 0.05 were included in our analysis.

#### Targeted fecal metabolome profiling

The absolute quantification of fecal samples (n = 1012) was performed by an ultra-performance liquid chromatography coupled to tandem mass spectrometry (UPLC-MS/MS) system. Briefly, the order of all samples was randomly selected prior to preparation. 10 mg lyophilized feces were homogenized with 25μL water and extracted with 185μL cold ACN-Methanol (8/2, v/v). At Biomek 4000 station

(Biomek 4000, Beckman Coulter, Inc., Brea, California, USA), 30µl centrifuged supernatant was derived with 20µl freshly prepared derivatives and mixed with internal standards in 30°C for 60min. The derivatization agents were 200 mM 3-NPH in 75% aqueous methanol and 96 mM EDC-6% pyridine solution in methanol. After derivatization, 350µl ice-cold 50% methanol solution was added to dilute the sample and then retained at -20°C for 20 minutes. After 4000g centrifugation at 4 °C for 30 minutes, 135µl supernatant was mixed and sealed with internal standards for each sample. Subsequently, the derivatized samples and serial dilutions of derivatized stock standards were analyzed randomly and quantitated by the UPLC-MS/MS. The instrument setting was: ACQUITY UPLC BEH C18 analytical column (2.1*100 mm,1.7µM); column temperature 40 °C; flow rate 0.4 mL/min; mobile phases A (water with 0.1% formic acid), mobile phases B (acetonitrile: IPA, 90:10); 0-1 min (5% B), 1-12 min (5-80% B), 12-15 min (80-95% B), 15-16 min (95-100%B), 16-18 min (100%B), 18-18.1 min (100-5% B), 18.1-20 min (5% B); 1.5Kv (ESI+), 2.0Kv (ESI-) capillary.

Three types of quality control samples, i.e. test mixtures, internal standards, and pooled biological samples were used in the metabolomics platform. The internal standards were added to the test samples in order to monitor analytical variations during the entire sample preparation and analysis process. The derivatized pooled samples for quality control were injected per14 samples. Raw data generated by UPLC-MS/MS were processed using the QuanMET software (v2.0, Metabo-Profile,

Co., Ltd, Shanghai, China) to perform peak integration, calibration, and quantification for each metabolite. The list of metabolites was selected to capture the microbiota-related metabolites and some key host metabolites. Finally, 117 metabolites were selected. These metabolites mainly include amino acids, bile acids and fatty acids.

**Statistical analysis**

All statistical analyses were performed using Stata version 15 or R version 4.0.2. Participants were categorized into three groups (healthy, prediabetes and T2D) based on their diabetes status. To explore the compositional variation of gut antibiotic resistome, we correlated 37 factors (including demography, physiology and dietary factors) to the ARG subtype distance matrix (Bray-Curtis) using permutational multivariate analysis of variance (PERMANOVA). We then used Pearson correlation analysis to examine the association between α-diversity of gut antibiotic resistome and microbial gene richness. The principle coordinates analysis (PCoA) on Bray-Curtis distance and PERMANOVA were performed to examine the structural differences of gut antibiotic resistome and gut microbiota among three different groups using the *adonis* function (permutations = 999). In addition, Procrustes analysis was performed to investigate the relationship between gut antibiotic resistome and gut microbiota, and the $p$ value was generated based on 999 permutations. We then examined the association between α-diversity indices of gut antibiotic resistome and gut microbiota and prevalent T2D using a logistic regression model, adjusted for potential confounders as follows: age, sex, body mass index

(BMI), physical activity, smoking status, drinking status, education attainment, household income level, Bristol stool scale and MGR. The α-diversity indices were z-score normalized before regression analysis.

To identify the markers of T2D, we used the least absolute shrinkage and selection operator (LASSO) regression model with 5 repeated 5-fold cross-validations based on the gut ARGs, microbial species and main covariates (age, sex, BMI, physical activity, smoking status, drinking status, education attainment, household income level, systolic blood pressure, diastolic blood pressure and Bristol stool scale). LASSO was implemented in the R package *glmnet* using a binomial response type for binary dependent variables (Non-T2D (healthy and prediabetes)/T2D, healthy/T2D, prediabetes/T2D)[7]. We assessed the predictive performance of the selected models by estimating the area under the receiver operation curve (AUC) for binary responses (alpha = 1; 100 lambda tested) (Fig S6). The selected value of 'lambda.min' was defined using cross-validation, the lambda controls the overall impact of LASSO. Then we merged the features with nonzero coefficients of the three models (Non-T2D/T2D, healthy/T2D, prediabetes/T2D) as markers of T2D progression.

Subsequently, the abundances of the markers were z score transformed. We used Kruskal-Wallis test and Mann-Whitney U test to examine the abundance differences of the marker ARGs and microbial species among healthy, prediabetes and T2D groups. The logistic regression was performed to investigate the odds ratio of the markers for risk of T2D after adjustment for age, sex, BMI, smoking status, drinking status, education attainment, income level and physical activity. Here, $p$ values were controlled by Benjamini-Hochberg method for multiple tests. FDR-corrected or raw $p$ values < 0.05 were considered to be significant.

***Genome-wide association analysis of T2D-related ARG features.*** To further examine the probability that ARG features increased the risk of T2D, GWAS for ARG α-diversity indices and T2D positively related ARG markers were conducted in 947 participants with both host genetic and metagenomics data. For the targeted ARG features, we used log transformation and z-score normalization to change the skewed distribution before GWAS analysis. A mixed linear model-based leave-one-chromosome-out association (MLMA-LOCO) analysis in GCTA was used to assess the association, fitting the first five genetic principal components of ancestry, age and sex as fixed effects and the effects of all the SNPs as random effects[8].

***One sample Mendelian randomization analysis.*** To test if ARG features were causally linked to T2D, the genetic variants used for one sample MR analysis were extracted from the GNHS study with a moderate cutoff of $p < 5 \times 10^{-5}$. The weighted polygenic risk score for each trait was constructed with the effect size from the additive model. The two-stage one-sample analysis was implemented to estimate the potential casual association. The first stage included a regression of the ARGs or α-diversity index on the polygenic risk score, adjusted for age at the time of stool sample collection, sex and the first five genetic principal components of ancestry. The second stage included a logistic regression of T2D using the prediction value constructed with the first stage regression, adjusted for age, sex and the first five genetic principal components of ancestry. Results were presented as odds ratio per 1-SD increase in polygenic risk score.

Based on the identified T2D-related ARGs, we constructed a diabetes-ARG score (DAS) as a new feature to represent the gut antibiotic resistome associated with T2D. We used the formula to compute DAS as follows:

$$\mathrm{DAS} = \Sigma_{1}^{\mathrm{n}}(OR_{\mathrm{i}} - 1) \times A_{i} \qquad (2)$$

Where n is the number of marker ARGs of T2D progression; $OR_i$ is the odds ratio of the i-th marker for risk of T2D; $A_i$ is the i-th normalized abundance (z score) of ARGs.

To test the reliability of DAS, we performed a logistic regression analysis to examine the cross-sectional association between DAS and T2D, adjusted for potential confounders. In addition, we assessed the cross-sectional correlation between DAS and glycemic traits, including fasting blood glucose, HbA1c, insulin and HOMA-IR (homeostatic model assessment of insulin resistance). The linear regression analysis was performed after adjustment for age, sex, BMI, smoking status, drinking status, education attainment, income level, physical activity. Considering that T2D related taxonomies of the gut microbiota may confound the above association, we also adjusted the Diabetes-Microbiota Score (DMS) constructed by the same method as

DAS. Moreover, the linear mixed models were used to examine the longitudinal association between DAS (baseline) and glycemic traits (repeat measure at baseline and follow-up visit) after excluding the baseline T2D cases, adjusted for age, sex, BMI, smoking status, drinking status, education attainment, income level, physical activity and DMS. In addition, we used a multivariable linear regression model to assess the cross-sectional association of the gut antibiotic resistome features (including DAS and α-diversity indices) with other cardiometabolic risk factors, including BMI, waist circumference, total cholesterol, triglycerides, HDL cholesterol, LDL cholesterol, TC/HDL ratio, systolic blood pressure and diastolic blood pressure. The dependent variables with skewed distribution were log-transformed before analysis (fasting blood glucose, insulin, HOMA-IR, TC/HDL-C and TG). The regression associations were expressed as the difference in cardiometabolic risk factors (in SD unit) per 1 SD difference in each gut antibiotic resistome feature.

As we only used the resistome information at baseline of the cohort, it might be that the gut antibiotic resistome would change over time. To address this concern, we performed a Procrustes analysis in 278 participants of the cohort with a median follow-up of 3.2 years. The fecal samples of these participants were collected twice (baseline and a follow-up visit).

***Network analysis of ARG-microbe associations.*** Spearman correlation analysis was performed to examine the associations between T2D-related ARGs and T2D-related gut microbial species, based on ‘Co-occurrence Network Analysis’ package (github.com/RichieJu520) [9]. To explore the underlying associations among T2D-related ARGs and all gut microbial species, we constructed a correlation matrix by calculating the pairwise Spearman correlation coefficients. A correlation between ARG-ARG, species-species, or ARG-species was considered significant if FDR-corrected $p < 0.05$. We further applied Gephi to visualize the correlations (Spearman's rho was $\geq 0.3$) in a network interface and explore its topological properties.

To fully explore the hidden deterministic (or non-random) co-occurrence patterns, we also computed the global co-occurrence associations between all the gut ARGs and microbial species identified. The observed (O%) and random incidences (R%) of co-occurrence correlation between two group entities (i.e., ARG and/or species) were statistically checked using the method as described previously [9,10]. Briefly, O% was calculated as the number of observed edges divided by total number of edges in the observed network, while R% was theoretically calculated by considering the frequencies of two group entities and assuming random association. Here co-occurrence patterns with Spearman's rho $\geqslant 0.6$, O% $\geqslant 1.0$ / R% $\geqslant 1.0$, and O/R $\geqslant 1.5$ / O/R $\leqslant 0.5$ were considered as significant difference.

We finally used Spearman correlation analysis to investigate the associations between gut antibiotic resistome features (DAS, *Multidrug_emrE*, *Vancomycin_vanX*,

*Quinolone_norB* and *MLS_ermX*) and 117 fecal metabolites. The concentrations of the metabolites were transformed to z-scores before analysis.

**Fig. S1. Flow diagram of participants' selection for the analyses of present study.**

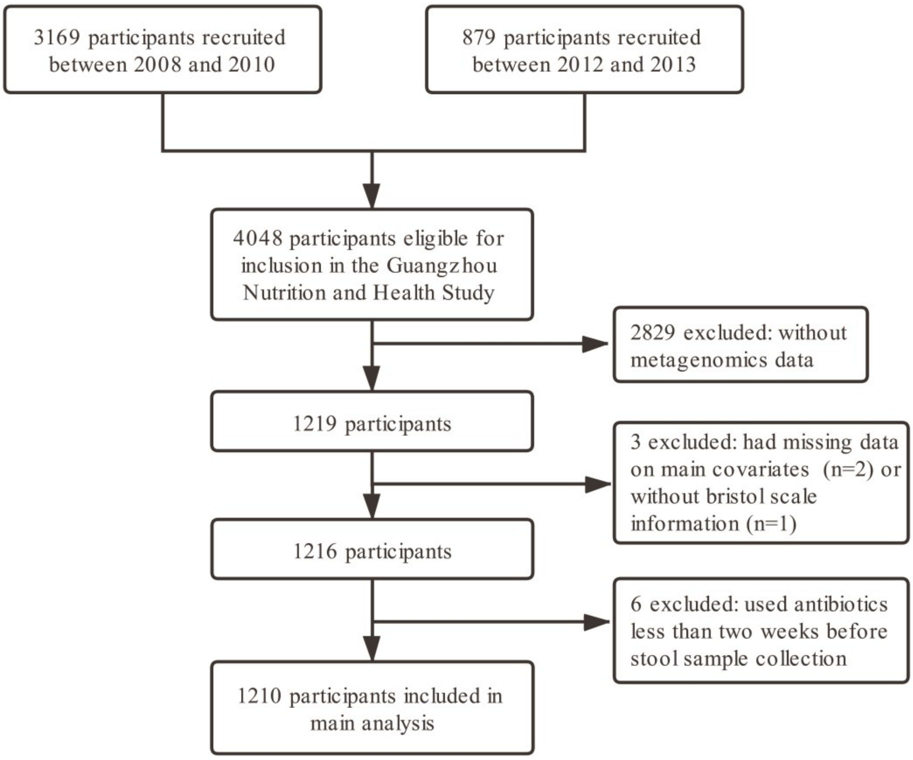

**Fig. S2. Workflow for analysis of metagenomics sequencing data.** Two distinct but complementary pipelines were used for metagenomics analysis. 19 antibiotic resistance gene (ARG) types and 805 ARG subtypes were annotated using ARG-OAP2. 639 microbial bacteria species were identified using MetaPhlAn2. PCoA, principal coordinates analysis.

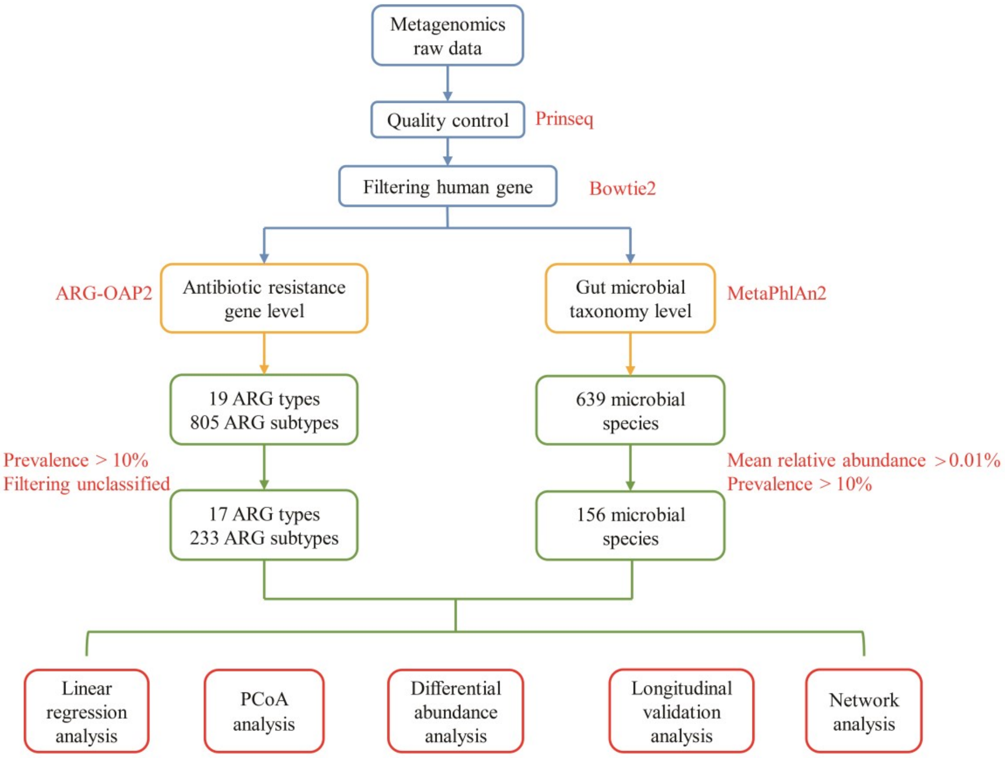

**Fig. S3. Abundance and prevalence of gut antibiotic resistome in GNHS. A**, The bar chart shows the prevalence of 17 antibiotic resistance genes (ARGs) types in participants with different diabetes status. **B**, The curve shows the association between ARGs subtype numbers and the prevalence among Healthy (n = 531), Prediabetes (n = 495) and T2D (n = 184) groups. **C**, The bar chart shows the prevalence of the ARGs types among different groups (differences between each two groups more than 3% are presented). **D**, The box plot shows the abundance of 17 core ARGs subtypes (prevalence = 100%). All box plots are the median with the interquartile range. *MLS*, *Macrolide-Lincosamide-Streptogramin*.

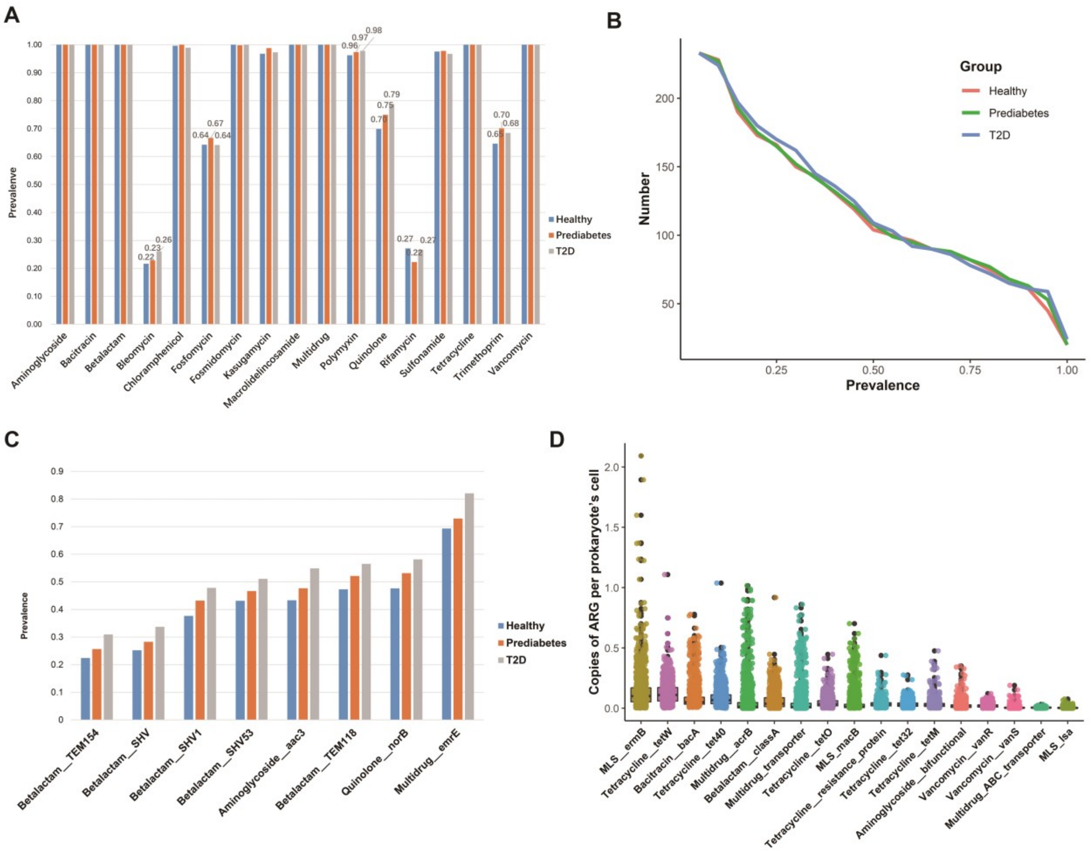

**Fig. S4. The effect sizes of host factors in gut antibiotic resistome grouped by diabetes status.** The effect sizes of host factors in human gut antibiotic resistome were calculated by PERMANOVA (Adonis, Bray-Curtis distance, permutations = 999) among Healthy (n = 392), Prediabetes (n = 401) and T2D (n = 154) groups.

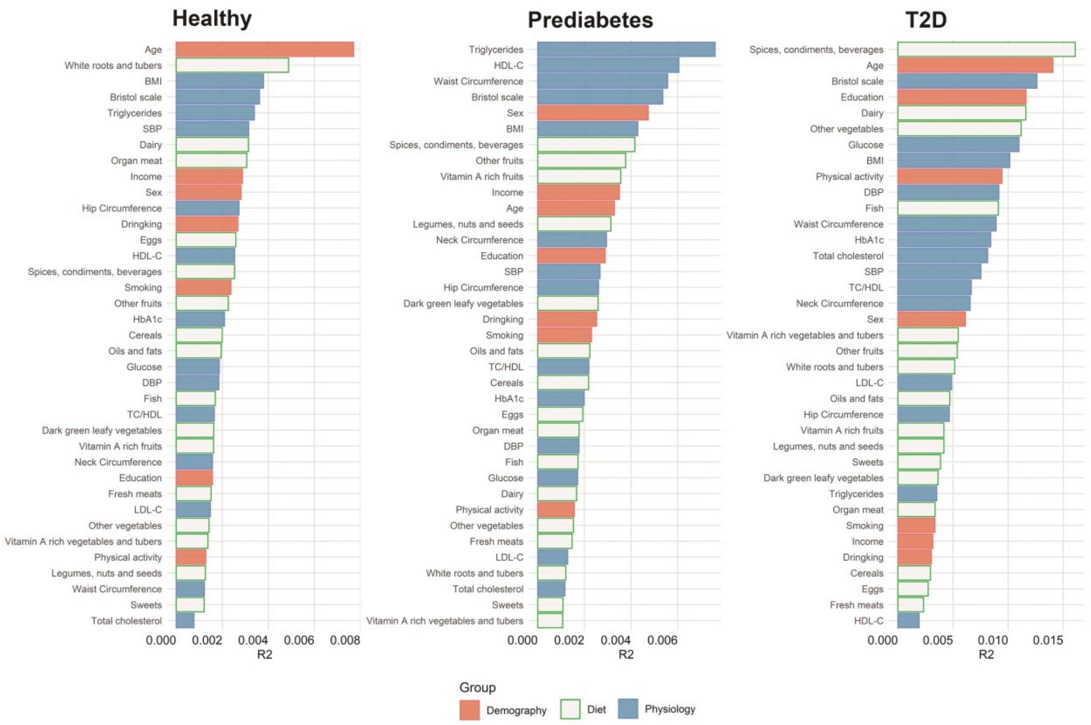

**Fig. S5. Correlations between gut antibiotic resistome diversity and microbiota.** **A**, **B,** Correlation between microbial gene richness (MGR) and α-diversity indices (ARGs) evaluated by Pearson tests. **C**, Procrustes analysis of gut ARGs versus gut microbiota. ARGs and microbiota are shown as orange and blue dots, respectively. ARGs and microbiota from the same individual are connected by grey lines.

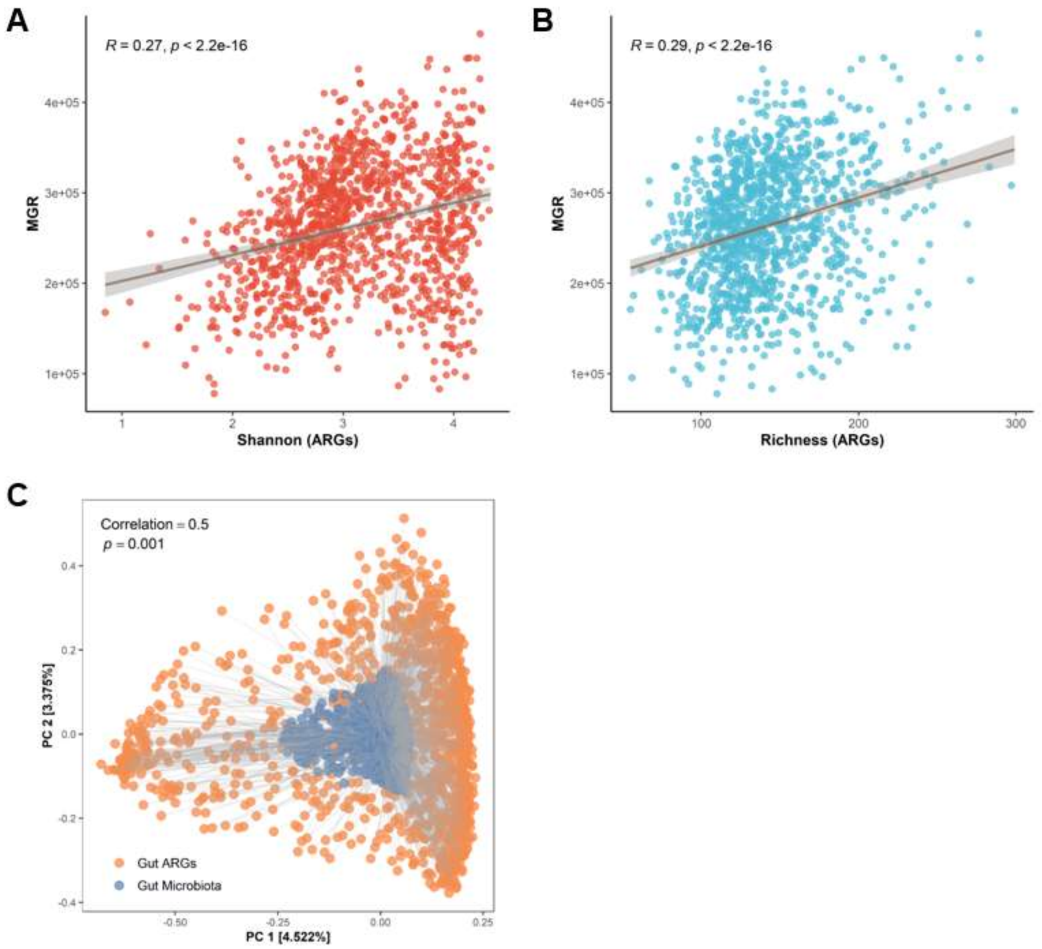

**Fig. S6. The comparison of inter-individual Bray-Curtis distance of gut antibiotic resistome.** Violin plots show the Bray-Curtis distance (y axis) among Healthy (n = 531), Prediabetes (n = 495) and T2D (n = 184) groups. *p* values from rank-based Wilcoxon test and Kruskal–Wallis test.

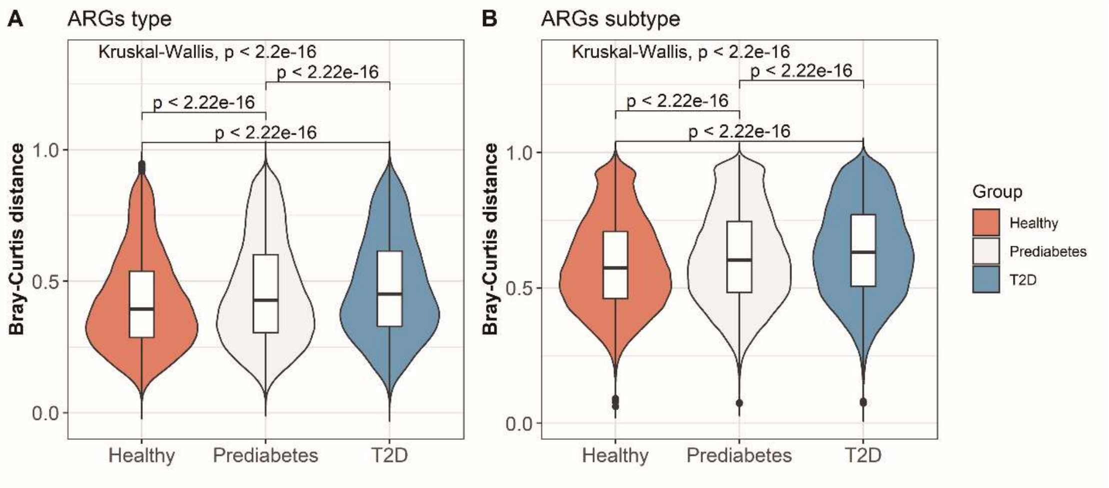

**Fig. S7. The performance of models based on LASSO feature selection.** LASSO regression models were performed with 5 repeated 5-fold cross-validations. The cross-validation AUCs were provided for both ARGs classifier (**A-C**) and microbiota classifier (**D-F**). Three dependent binary variables for antibiotic resistance genes marker selection: (**A**, **D**) Non-T2D (Healthy and Prediabetes)/T2D, (**B**, **E**) Healthy/T2D, (**C**, **F**) Prediabetes/T2D. LASSO, least absolute shrinkage and selection operator.

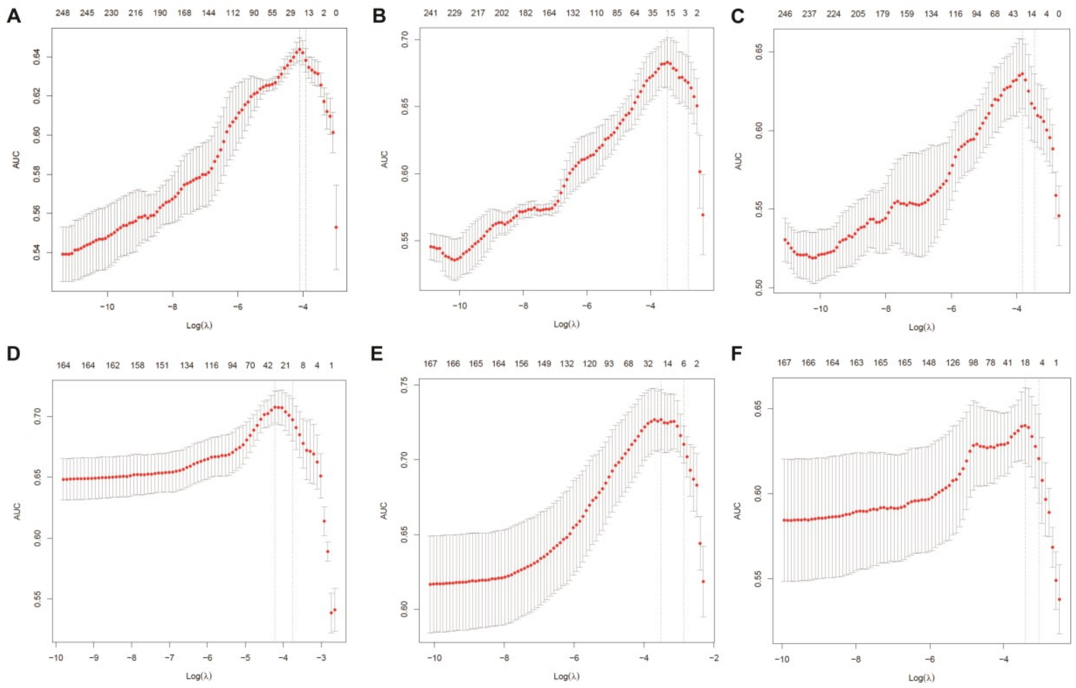

**Fig. S8. Venn plot of biomarkers identified by LASSO models.** Venn plot showing the number of biomarkers identified by gut ARGs classifier (**A**) and microbiota classifier (**B**) for different datasets: Non-T2D/T2D, Healthy/T2D and Prediabetes/T2D. LASSO, least absolute shrinkage and selection operator.

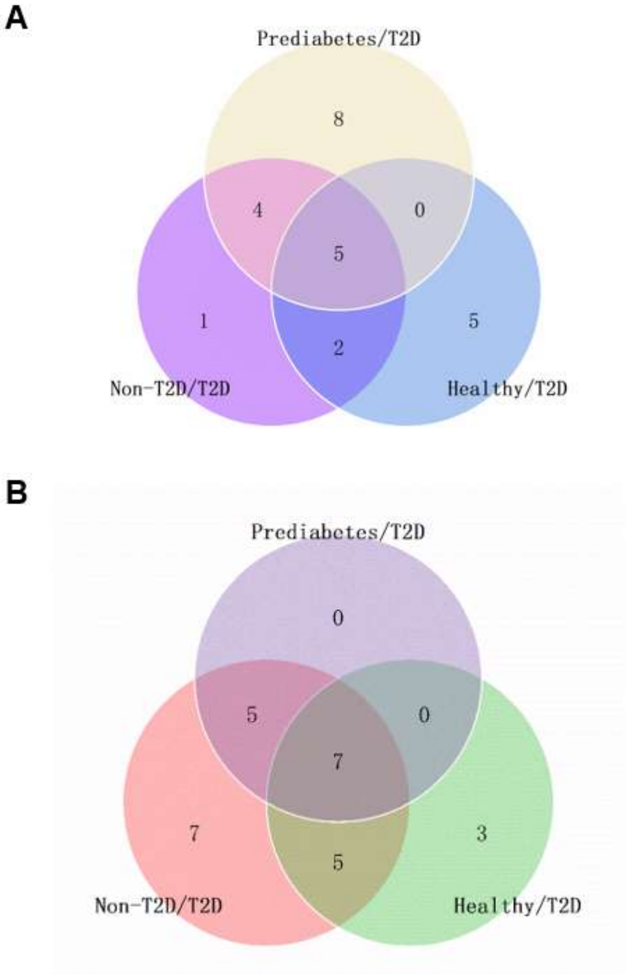

**Fig. S9. Networks of co-occurring T2D-related ARGs and gut microbiota, grouped by diabetes status.** Networks were presented based on correlation analysis among Healthy (n = 531), Prediabetes (n = 495) and T2D (n = 184) groups. A node stands for an ARG type/subtype or a species and a connection (i.e. edge) stands for a significant (FDR-corrected *p* < 0.05, Spearman's rho ≥ 0.3) pairwise correlation. Network was colored by ARGs and phylums. Node size is proportional to the number of connections (i.e. degree). *Ami*, *Aminoglycoside*; *Bet*, *Betalactam*; *Chl*, *Chloramphenicol*; *MLS*, *Macrolide-Lincosamide-Streptogramin*; *Mul*, *Multidrug*; *Qui*, *Quinolone*; *Tet*, *Tetracycline*; *Van*, *Vancomycin*.

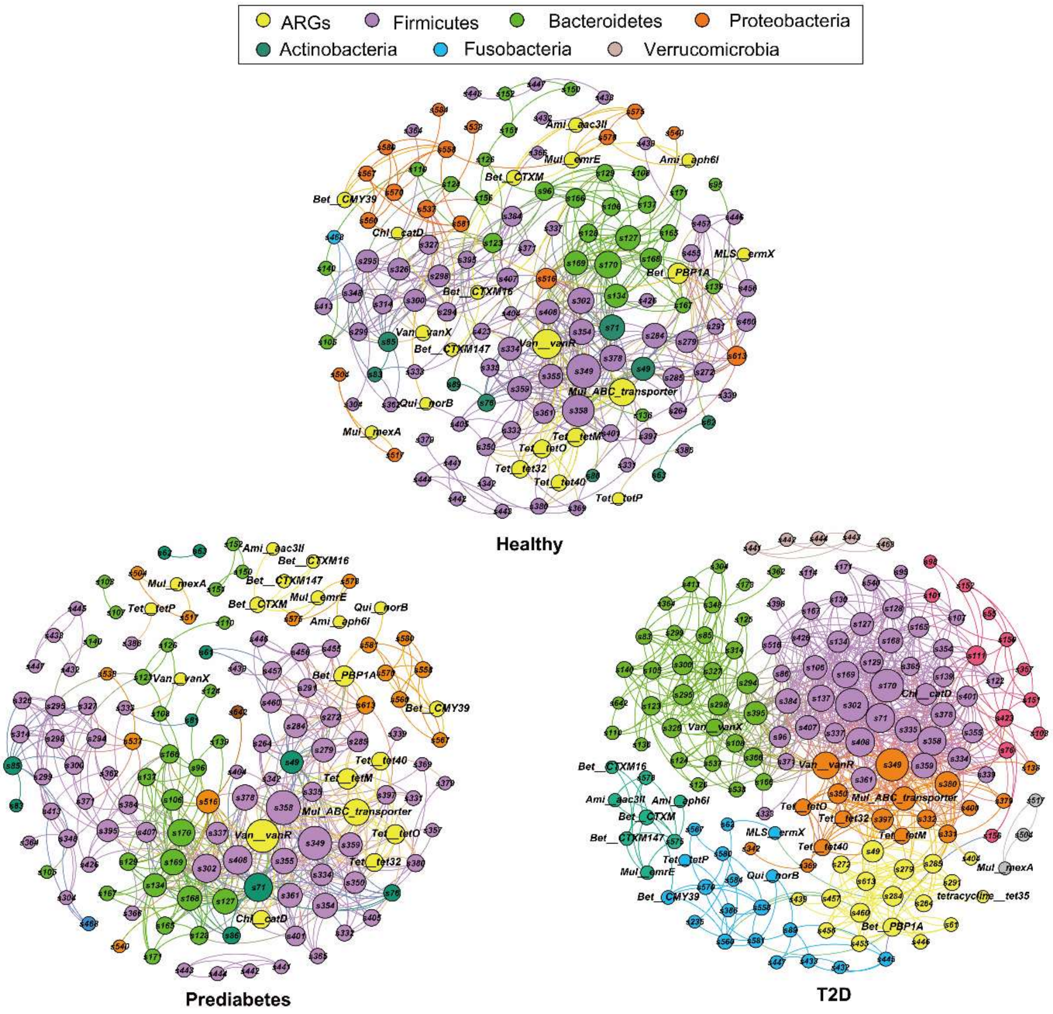

**Fig. S10. Associations between gut antibiotic resistome features and fecal metabolites (n = 1012).** The heatmap shows the Spearman correlation coefficients between gut antibiotic resistome features and fecal metabolites (purple text, showing fatty acids; yellow text, showing bile acids; blue text, showing amino acids). DAS, Diabetes-ARG score. *FDR-corrected $p < 0.05$, ** FDR-corrected $p < 0.01$, *** FDR-corrected $p < 0.001$.

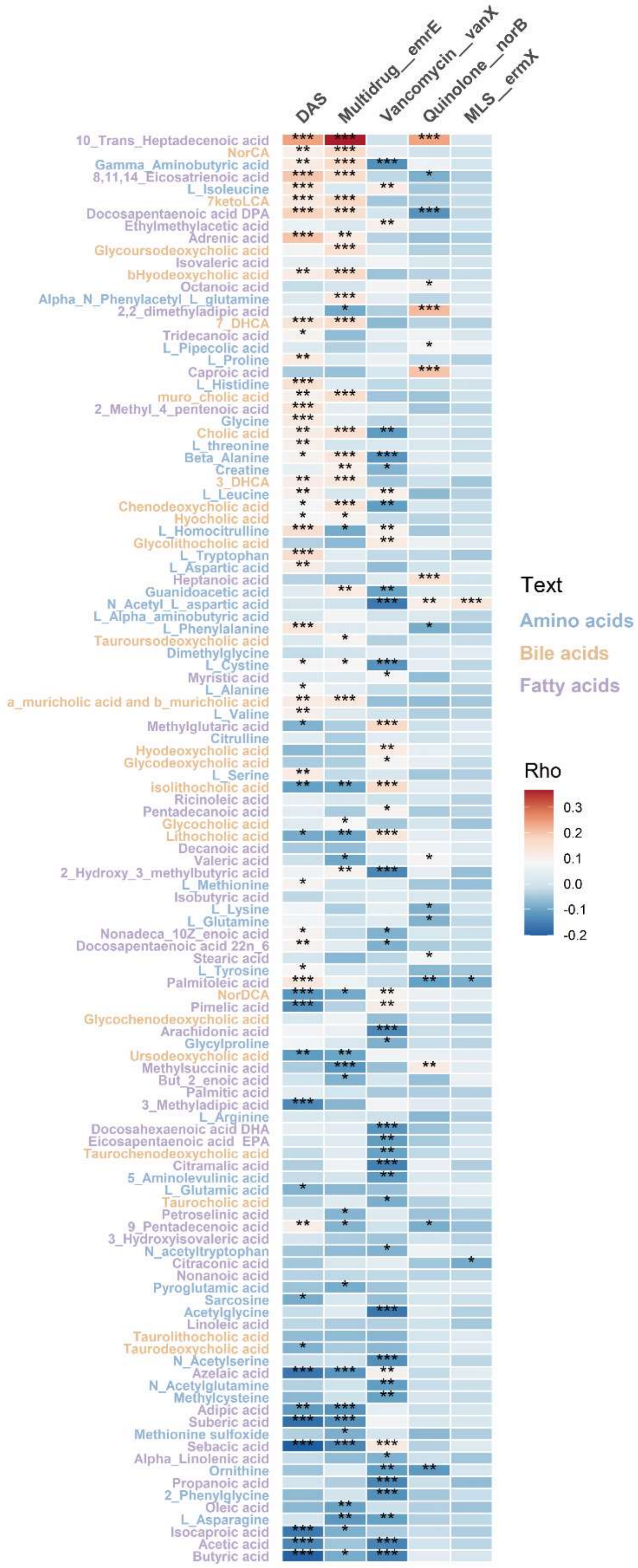
